# Screening for Asymptomatic Tuberculosis among Adults with Household Exposure to a Patient with Pulmonary Tuberculosis

**DOI:** 10.1101/2025.01.20.25320843

**Authors:** Simon C Mendelsohn, Humphrey Mulenga, Michele Tameris, Tumelo Moloantoa, Stephanus T Malherbe, Austin Katona, Fernanda Maruri, Firdows Noor, Ravindre Panchia, Khuthadzo Hlongwane, Kim Stanley, Yuri F van der Heijden, Katie Hadley, Dominique T Ariefdien, Novel N Chegou, Gerhard Walzl, Thomas J Scriba, Timothy R Sterling, Mark Hatherill, the RePORT South Africa Study Team

**Affiliations:** South African Tuberculosis Vaccine Initiative, Institute of Infectious Disease and Molecular Medicine and Division of Immunology, Department of Pathology, University of Cape Town, Cape Town, South Africa; Perinatal HIV Research Unit, University of Witwatersrand, Johannesburg, South Africa; DST/NRF Centre of Excellence for Biomedical TB Research; South African Medical Research Council Centre for TB Research; Division of Immunology, Department of Biomedical Sciences, Faculty of Medicine and Health Sciences, Stellenbosch University, Cape Town, South Africa; Vanderbilt Tuberculosis Center, Vanderbilt University Medical Center, Nashville, TN, USA; The Aurum Institute, Johannesburg, South Africa

**Keywords:** asymptomatic, tuberculosis, household, contacts, active case finding, screening

## Abstract

**Background:** More than half of tuberculosis (TB) detected by community prevalence surveys is classified as asymptomatic. We evaluated yield of symptom and chest radiograph (CXR) screening of TB-exposed household contacts (HHC) in South Africa.

**Methods:** Adult volunteers (≥18 years) with household exposure to pulmonary TB patients were enrolled at three sites. Systematic screening of TB symptoms (any duration), CXR (any abnormality), and sputum microscopy, Xpert Ultra, and liquid culture were performed. Serum C-reactive protein (CRP) was measured by multiplex bead array. Prevalent TB was microbiologically-confirmed (Xpert Ultra or culture). Symptomatic and asymptomatic TB were defined as prevalent TB with and without reported symptoms, respectively.

**Results:** Between March 2021 – December 2022, 979 HHC were enrolled; 185 (18.9%) living with HIV and 187 (19.1%) with previous TB. Prevalent TB occurred in 51 (5.2%) and was asymptomatic in 42/51 (82.4%). Only 13/42 (31.0%) asymptomatic TB cases were smear-positive [8/13 (61.5%) graded scanty or 1+]. CRP did not discriminate healthy HHC from those with asymptomatic TB (AUC 0.60; 95%CI 0.47–0.73). An abnormal CXR was observed in 23/41 asymptomatic (sensitivity 56.1%, 95%CI 41.0–70.1%) versus 8/9 symptomatic (sensitivity 88.9%, 95%CI 56.5–98.0%) TB cases. Sensitivity of CXR in combination with symptom screening was 64.0% (32/50, 95%CI 50.1–75.9%) for all prevalent TB.

**Conclusions:** More than 80% of confirmed TB cases among HHC were asymptomatic. CXR screening missed more than 40% of these asymptomatic cases. Community prevalence surveys reliant on symptom- and CXR-based approaches may significantly underestimate the prevalence of asymptomatic TB in endemic countries.

**Funding:** Supported by RePORT South Africa through funding from the U.S. National Institutes of Health, CRDF Global, and the South African Medical Research Council.

**RESEARCH IN CONTEXT:** *Evidence before this study:* World Health Organisation (WHO) guidelines for systematic tuberculosis (TB) screening recommend symptom screening and chest radiography (CXR), based on a Cochrane meta-analysis reporting 70.6% sensitivity (any TB symptom) and 94.7% sensitivity (any CXR abnormality) for bacteriologically-confirmed pulmonary TB. National TB prevalence surveys rely on a positive symptom screen or abnormal CXR to trigger diagnostic sputum testing. This approach to community screening would, by definition, miss asymptomatic TB cases without CXR evidence of disease. We reviewed the reference list of the aforementioned meta-analysis for active case-finding studies of adolescents and adults aged 15 years and older in community and contact-tracing settings. We performed forward citation-tracking and searched reference lists, including studies published in English between Jan 1, 1980, and November 1, 2024. We excluded studies that included children <15 years; or that exclusively enrolled people with additional risk factors (HIV; diabetes; latent TB infection; prior TB). We found 28 studies that performed universal sputum testing for bacteriologically-confirmed pulmonary TB and reported 51.8% (95%CI 49.9–53.7%; *I*^2^ = 89.2%) pooled sensitivity for symptom screening (any symptom; 24 studies, 2,969 TB cases) and 62.4% (95%CI 59.3–65.3%; *I^2^* = 88.3%) pooled sensitivity for CXR (any abnormality; 10 studies, 1,123 TB cases). Only four studies (145 TB cases) reported accuracy of symptom screening in parallel with chest radiography (pooled sensitivity 67.3%, 95%CI 57.3–75.9%; *I^2^* = 87.1%), but these studies did not disaggregate symptomatic and asymptomatic disease.

*Added value of this study:* We performed systematic screening using universal sputum microbiological testing of 978 household contacts of pulmonary TB patients in three South African communities and compared symptom (any duration) and CXR (any abnormality) screening approaches against a microbiological reference standard. We detected confirmed pulmonary TB in 5.2% of household contacts, and 82.4% of these TB cases reported no TB symptoms. Asymptomatic TB in household contacts was pauci-bacillary and associated with low serum CRP levels that were indistinguishable from healthy controls, but distinct from symptomatic TB in a comparator group of clinic attendees. Sensitivity of CXR screening for asymptomatic TB was only 56.1%; sensitivity of combined symptom and CXR screening for all TB was marginally higher at 64.0%.

*Implications of all the available evidence:* Our findings from household contacts suggest that symptom- and CXR-based approaches are inadequate for community TB screening in South Africa and do not meet the WHO Target Product Profile for a TB screening test (minimum 90% sensitivity; 70% specificity). National TB Prevalence Surveys that omit universal sputum microbiological testing may significantly underestimate the prevalence of asymptomatic TB in high-burden countries.

## INTRODUCTION

Approximately 2.7 million (25%) of the estimated 10.8 million global tuberculosis (TB) cases went undiagnosed or untreated in 2023.^1^ To decrease *Mycobacterium tuberculosis* (*Mtb*) transmission and reduce the global burden of TB disease, it is necessary to find and treat these so-called “missing millions”. However, more than half of all TB found in community TB prevalence surveys has been classified as asymptomatic^2,3^—occurring in persons who do not have, recognise, or report typical TB symptoms such as cough, fever, night sweats, and loss of weight.^4^

The World Health Organisation (WHO) has recently recognised the importance of asymptomatic (previously “subclinical”) TB for disease transmission and is currently reviewing its guidance to determine programmatic implications.^5^ In mathematical models an estimated 50% of asymptomatic TB cases will never develop symptoms and recover, but left untreated, the other 50% remain infectious, and at risk of progression to symptomatic TB disease and death.^6^ Despite purportedly being less infectious, epidemiological and modelling studies suggest that asymptomatic TB is responsible for a greater share of transmission than symptomatic disease, due to longer duration of exposure before treatment.^7–9^

It follows that finding and treating asymptomatic TB in the community is important, but, by definition, individuals with asymptomatic TB would not be detected by symptom-triggered TB surveillance. New tools are needed for community TB screening. Since ascertainment of symptoms is subjective, health-seeking behaviour may be a more important determinant of diagnostic performance than simply the presence or absence of symptoms. It remains to be shown whether tools developed for triage of clinic attendees with symptomatic TB would perform similarly for community screening of asymptomatic TB, which is likely less severe.

Most active case-finding studies, prevalence surveys, and public health programmes rely on symptom and/or chest radiographic (CXR) screening,^2,3,10^ to prioritise resources for diagnostic sputum testing. World Health Organisation (WHO) guidelines^11^ also recommend symptom and CXR screening, based on a Cochrane meta-analysis reporting 70.6% sensitivity (any TB symptom) and 94.7% sensitivity (any CXR abnormality) for bacteriologically-confirmed pulmonary TB.^10^ However, these data are primarily derived from national TB prevalence surveys, which rely on a positive symptom screen or abnormal CXR to trigger diagnostic sputum testing, and would miss asymptomatic TB cases without CXR evidence of disease.

Review of evidence before this study showed 51.8% (95%CI 49.9–53.7%; *I*^2^ = 89.2%) pooled sensitivity for symptom screening (any symptom; 24 studies, 2,969 TB cases) and 62.4% (95%CI 59.3–65.3%; *I*^2^ = 88.3%) pooled sensitivity for CXR (any abnormality; 10 studies, 1,123 TB cases) in studies that performed universal sputum testing for bacteriologically-confirmed pulmonary TB (references in **Supplement**). Only four studies (145 TB cases) reported accuracy of symptom screening in parallel with chest radiography (pooled sensitivity 67.3%, 95%CI 57.3–75.9%; *I*^2^ = 87.1%), but these studies did not disaggregate symptomatic and asymptomatic disease (**Figure 1**).

We aimed to understand the burden of asymptomatic TB disease among TB-exposed household contacts (HHC) in communities in South Africa; and to evaluate the yield of systematic screening using universal sputum microbiological testing, symptomatology, and CXR.

**Figure 1.**
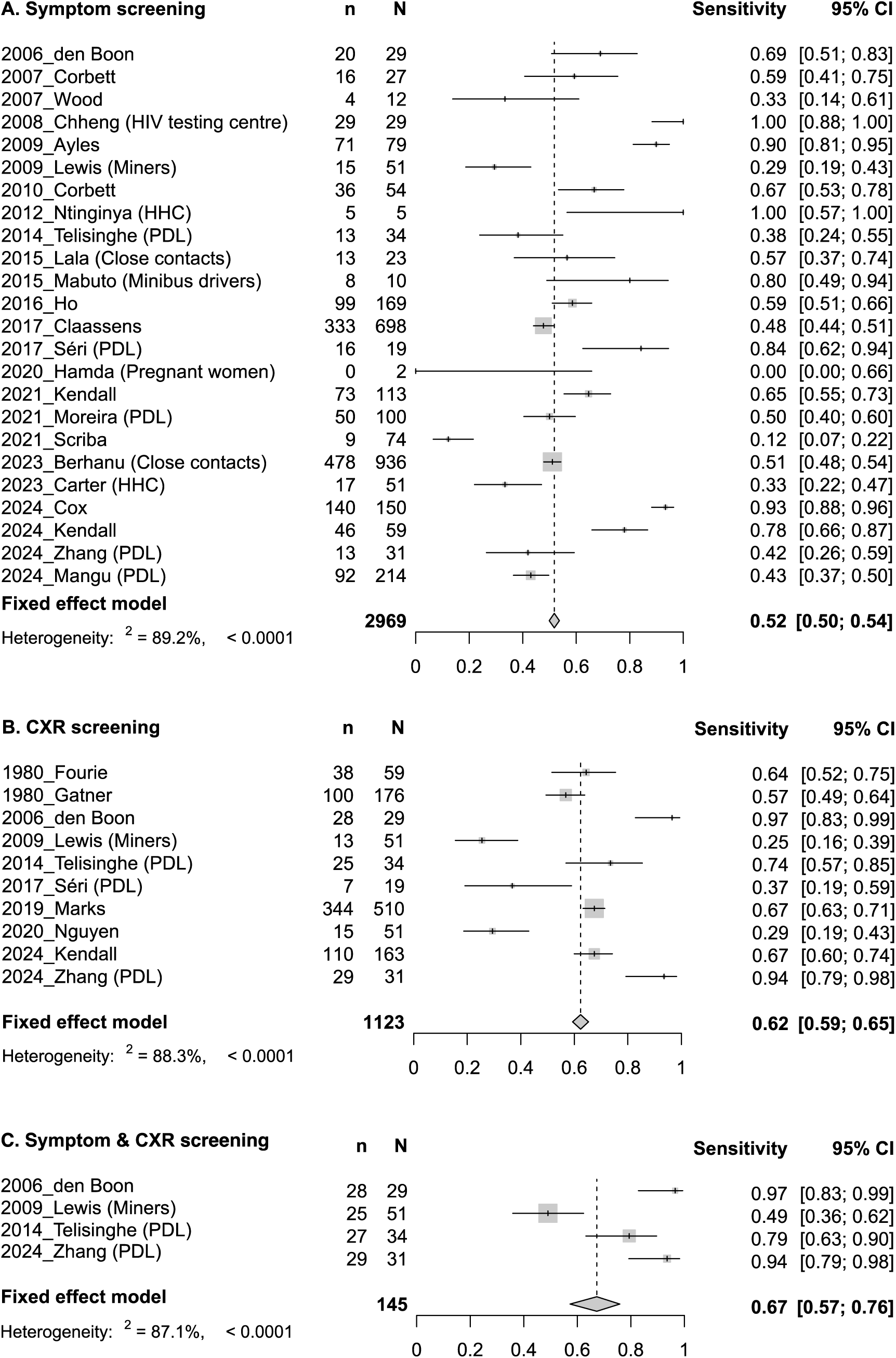
Sensitivity of symptom and chest radiography (CXR) screening for tuberculosis (TB) with universal sputum microbiological testing irrespective of presence of symptoms or CXR abnormality. Forest plot of sensitivity of (A) symptom screening, (B) CXR, and (C) parallel symptom and CXR screening for TB reported in the literature. Cohorts are listed by year of publication and first author. HHC, household contacts. PDL, people deprived of liberty.

## METHODS

### Study design and participants

The Regional Prospective Observational Research for Tuberculosis (RePORT) South Africa network enrolled participants in a prospective observational cohort study at three centres in South Africa (Worcester and Ravensmead, Western Cape Province; Soweto, Gauteng Province). Adults (≥18 years) had recent household exposure within the past six months to an adult index case with untreated or inadequately treated pulmonary TB. Exposure was defined as sleeping in the same household, or more than four hours of other household exposure per week. Participants were excluded if they were unlikely to attend study visits, or had any condition that might interfere with their ability to provide informed consent or adhere to study requirements, including alcohol or drug dependence and incarceration.

Symptom screening, CXR, and spontaneously expectorated sputum collection for Xpert Ultra (Cepheid, CA, USA), liquid culture (Mycobacteria Growth Indicator Tube [MGIT], BACTEC, Beckton Dickinson, NJ, USA), and acid-fast microscopy, were performed at a baseline visit for all HHC, regardless of symptoms. A participant was classified symptomatic if they reported one or more of cough, fever, weight loss, fatigue, night sweats, pleuritic chest pain, or haemoptysis, for any duration. CXR were read once by an investigator at each site using a standardised form and recorded as normal or abnormal, based on absence or presence of any cavitation, opacity, mediastinal or hilar adenopathy, pleural effusion, bronchiectasis, or collapsed lung.

The microbiological reference standard (MRS) for microbiologically-confirmed pulmonary TB disease was at least one sputum specimen positive by MGIT culture or Xpert Ultra (excluding trace positive results). Sputum induction was not performed as it is not the current standard of care in South Africa. Participants who were unproductive of sputum, or with Xpert Ultra trace positive result only, were considered sputum-negative in the primary analysis. Participants who, after investigation for TB at baseline, were not confirmed by positive sputum MGIT culture or Xpert Ultra, were defined as controls for the purpose of diagnostic analyses. The definition of prevalent TB was restricted to cases diagnosed on sputum samples collected at the baseline visit. Symptomatic and asymptomatic TB were defined as microbiologically-confirmed prevalent TB with or without reported symptoms of any duration, respectively. All participants diagnosed with TB disease were referred for treatment.

### Measurement of C-reactive protein (CRP)

CRP was measured in HHC, and in a cohort of symptomatic clinic attendees recruited contemporaneously by the RePORT South Africa network (described in the **Supplement**). This exploratory analysis aimed to understand interactions between symptomatology and health-seeking behaviour. CRP was measured using a multiplex bead array in cryopreserved serum samples to differentiate asymptomatic TB from asymptomatic controls among HHC; and symptomatic TB from symptomatic controls among clinic attendees. Briefly, serum samples were collected at baseline, prior to TB diagnosis, and cryopreserved in 607 adults (≥18 years) self-presenting to clinics with presumptive TB (199 sputum microbiologically-confirmed symptomatic pulmonary TB; 408 symptomatic controls with negative sputum culture and Xpert Ultra) at five sites in South Africa. Samples were later thawed and assayed using multiplex bead array (Bio-Plex Pro Human Apolipoprotein 10-plex Assay Panel, BioRad) on the Bio Plex platform (Bio Plex, Bio Rad Laboratories, CA, USA) as previously described.^12^

### Statistical analysis

Analyses were performed using Stata (version 16.1, StataCorp, TX, USA) and R (version 4.4.1). Sensitivity, specificity, and area under the curve (AUC) were calculated using standard methods. Confidence intervals for diagnostic accuracy measures were calculated with the Wilson method and DeLong method for AUC. The number needed to test (NNT) with a confirmatory test to diagnose one TB case is calculated as the number of confirmatory tests divided by the number of true positives with the screening approach. CRP concentrations were censored at 150 mg/dL.

### Ethical approval

The study protocol was approved by the institutional research ethics committees at each participating South African site and at Vanderbilt University Medical Center, USA. All participants provided written, informed consent prior to participation.

## RESULTS

### Recruitment and baseline status of participants

Between March 2021 and December 2022, 1,001 HHC were screened and 979 enrolled (**Figure 2**). Most common reasons for exclusion included insufficient exposure to an index TB case (n=7), active psychiatric condition, or alcohol or drug dependence (n=5), and other miscellaneous reasons (n=10; **Figure 2**). Of the 979 HHC enrolled, 345 (35.2%) were male, median age was 34.8 years (interquartile range, IQR 25.4–48.2), 185 (18.9%) were living with HIV, 187 (19.1%) had known previous TB, and 834 (85.2%) were asymptomatic (**Table 1**).

Most participants (962/979; 98.3%) were able to provide a spontaneous expectorated sputum sample for testing. Prevalent, microbiologically-confirmed, pulmonary TB disease was diagnosed in 51/979 (5.2%) participants: 30 (58.8%) by sputum Xpert Ultra and MGIT culture, 13 (25.5%) by sputum MGIT culture alone, and 8 (15.7%) by sputum Xpert Ultra alone. Among those diagnosed by Xpert Ultra alone, 62.5% (5/8) had a previous TB episode, two of which occurred within the preceding three years (1.4 and 1.9 years).

**Figure 2.**
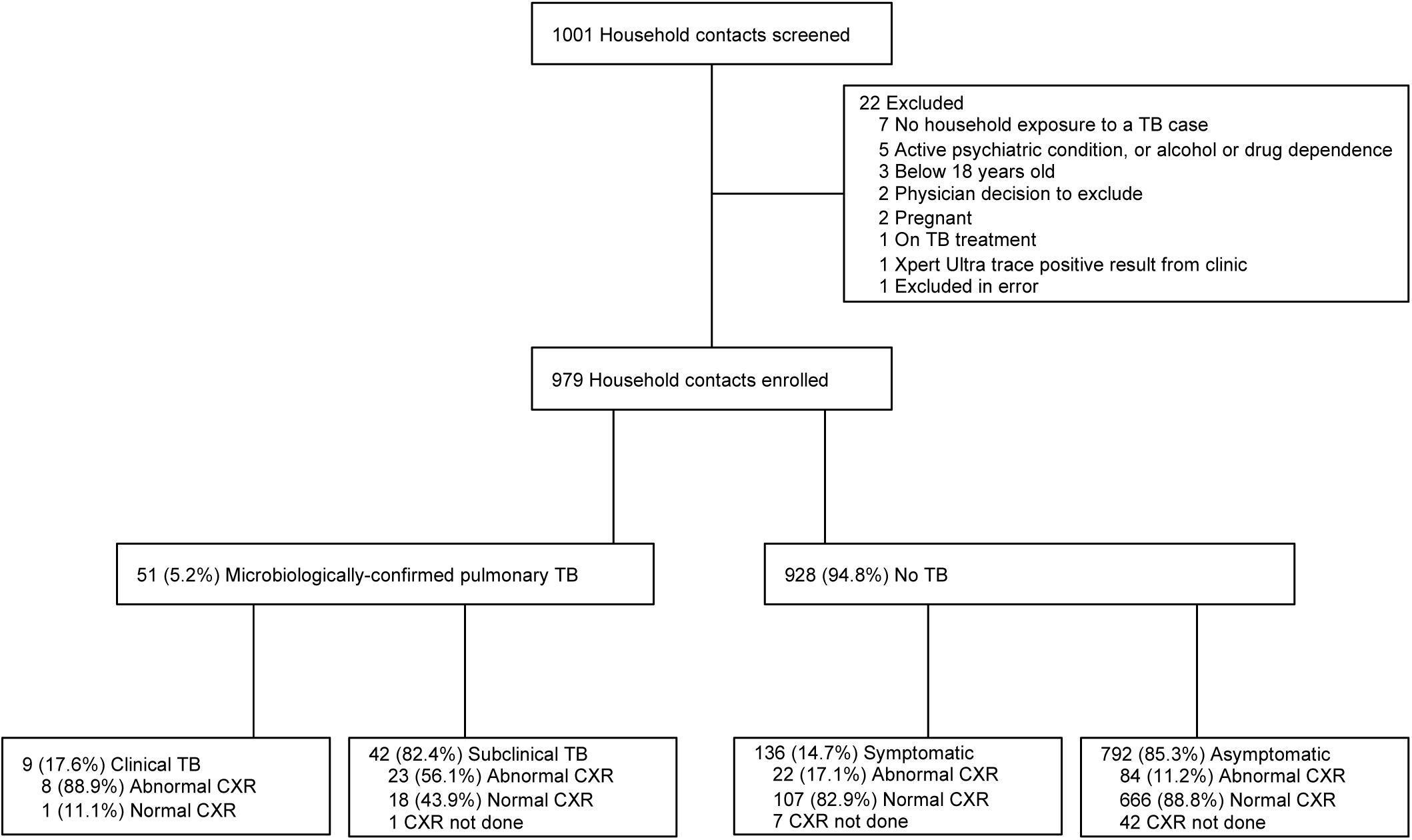
Study flow diagram. TB, tuberculosis. CXR, Chest radiograph.

**Table 1.** Characteristics of the study population by TB phenotype.

|  | Total<br>(N=979) | Asymptomatic<br>HHC with TB<br>disease<br>(N=42) | Symptomatic<br>HHC with TB<br>disease<br>(N=9) | Asymptomatic<br>HHC without TB<br>(N=792) | Symptomatic<br>HHC without TB<br>(N=136) |
| --- | --- | --- | --- | --- | --- |
| Median age (IQR) | 34.8 (25.4–48.2) | 35.8 (25.8–49.3) | 40.3 (31.7–49.6) | 33.3 (24.7–45.8) | 43.4 (32.0–53.8) |
| Male sex, n (%) | 345 (35.2) | 21 (50.0) | 4 (44.4) | 269 (34.0) | 51 (37.5) |
| Ancestry, n (%) |  |  |  |  |  |
| Black African | 463 (47.3) | 6 (14.3) | 3 (33.3) | 349 (44.1) | 105 (77.2) |
| Mixed ancestry | 512 (52.3) | 36 (85.7) | 6 (66.7) | 439 (55.4) | 31 (22.8) |
| Caucasian | 4 (0.4) | 0 | 0 | 4 (0.5) | 0 |
| History of smoking, n (%) |  |  |  |  |  |
| Never | 410 (41.9) | 10 (23.8) | 1 (11.1) | 340 (43.0) | 59 (43.4) |
| Current smoker | 515 (52.6) | 30 (71.4) | 6 (66.7) | 414 (52.3) | 65 (47.8) |
| Former smoker | 54 (5.5) | 2 (4.8) | 2 (22.2) | 38 (4.8) | 12 (8.8) |
| History of drug use, n (%) | 129 (13.2) | 11 (26.2) | 3 (33.3) | 98 (12.4) | 17 (12.5) |
| Prior TB, n (%) | 187 (19.1) | 13 (31.0) | 4 (44.4) | 134 (16.9) | 36 (26.5) |
| HIV positive, n (%) | 185 (18.9) | 7 (16.7) | 2 (22.2) | 129 (16.3) | 47 (34.6) |
| Chest radiography, n (%) |  |  |  |  |  |
| Normal | 792/929 (85.3) | 18/41 (43.9) | 1/9 (11.1) | 666/750 (88.8) | 107/129 (82.9) |
| Any abnormality suggestive of TB | 137/929 (14.7) | 23/41 (56.1) | 8/9 (88.9) | 84/750 (11.2) | 22/129 (17.1) |
| Not done | 50 | 1 | 0 | 42 | 7 |
| Median persons in household (IQR) | 6 (4–8) | 6 (4–9) | 6 (3–8) | 6 (4–8) | 5 (4–7) |
| Study site, n (%) |  |  |  |  |  |
| Ravensmead | 167 (17.1) | 6 (14.3) | 0 (0) | 160 (20.2) | 1 (0.7) |
| Worcester | 447 (45.7) | 29 (69.0) | 6 (66.7) | 382 (48.2) | 30 (22.1) |
| Soweto | 365 (37.3) | 7 (16.7) | 3 (33.3) | 250 (31.6) | 105 (77.2) |
| TB symptoms, n (%) | 145 (14.8) | 0 | 9 (100) | 0 | 136 (100) |
| Chest pain | 35 (3.6) | 0 | 4 (44.4) | 0 | 31 (22.8) |
| Cough | 104 (10.6) | 0 | 8 (88.9) | 0 | 96 (70.6) |
| Fever | 36 (3.7) | 0 | 1 (11.1) | 0 | 35 (25.7) |
| Fatigue | 24 (2.5) | 0 | 5 (55.6) | 0 | 19 (14.0) |
| Loss of weight | 57 (5.8) | 0 | 6 (66.7) | 0 | 51 (37.5) |
| Night sweats | 39 (4.0) | 0 | 6 (66.7) | 0 | 33 (24.3) |
| Microbiological confirmation, n (%) |  |  |  |  |  |
| Smear positive | 24 (2.5) | 13 (31.0) | 4 (44.4) | 5 (0.6) <sup>a</sup> | 2 (1.5) <sup>a</sup> |
| Scanty or 1+ | 15 (1.5) | 8 (19.0) | 1 (11.1) | 4 (0.5) <sup>a</sup> | 2 (1.5) <sup>a</sup> |
| 2+ | 7 (0.7) | 4 (9.5) | 2 (22.2) | 1 (0.1) <sup>a</sup> | - |
| 3+ | 2 (0.2) | 1 (2.4) | 1 (11.1) | - | - |
| Culture positive | 43 (4.4) | 35 (83.3) | 8 (88.9) | - | - |
| Time to positivity (days) | 14 (7–18) | 14 (7–17) | 12 (9–18) | - | - |
| Xpert Ultra positive | 47 (4.8) | 35 (83.3) | 9 (100) | 3 (0.4) <sup>a</sup> | 0 |
| Trace <sup>a</sup> | 9 (0.9) | 5 (11.9) | 1 (11.1) | 3 (0.4) <sup>a</sup> | 0 |
| Very low | 12 (1.2) | 10 (23.8) | 2 (22.2) | - | - |
| Low | 10 (1.0) | 8 (19.0) | 2 (22.2) | - | - |
| Medium | 6 (0.6) | 4 (9.5) | 2 (22.2) | - | - |
| High | 10 (1.0) | 8 (19.0) | 2 (22.2) | - | - |

<sup>a</sup> Reference standard for above analysis is positive sputum culture or Ultra, excluding trace positive. HHC, household contact. IQR, interquartile range. TB, tuberculosis.

### Most TB diagnosed among household contacts was asymptomatic

The majority (42/51; 82.4%, 95%CI 69.7–90.4%) of prevalent TB cases did not report TB symptoms of any duration and were classified as asymptomatic. Among asymptomatic TB cases, 31.0% (13/42) reported a previous TB episode, seven of which occurred within the preceding three years, and 76.2% (32/42) were current or former smokers.

CXR was performed in 97.6% (41/42) of asymptomatic TB cases, among whom an abnormal CXR was observed in 56.1% (23/41; 95%CI 41.0–70.1%); of whom 39.1% (9/23) reported a prior TB history. Conversely, an abnormal CXR was observed in 88.9% (8/9; 95%CI 56.5–98.0%) of symptomatic TB cases (difference in proportions 32.8%, 95%CI -2.5–50.4%); and 12.1% (106/879; 95%CI 10.1–14.4%) of all controls without TB. No CXR abnormality was observed in 43.9% (18/41; 95%CI 29.9–59.0%) of asymptomatic TB cases.

### Asymptomatic TB was associated with low sputum bacillary load

Only 13/42 (31.0%) asymptomatic TB cases were sputum smear microscopy positive; most (8/13, 61.5%) were graded scanty or 1+. Similarly, most of the 35/42 (83.3%) Xpert Ultra positive asymptomatic TB cases occurred in the “trace” (not included in MRS) and “very low” semi-quantitative categories (15/35, 42.9%), with 8 (22.9%), 4 (11.4%), and 8 (22.9%) additional cases graded “low”, “medium”, and “high”, respectively. Median time-to-culture-positivity for asymptomatic TB cases was 14 (IQR 7–17) days.

Symptomatic TB cases had a slightly higher rate of sputum smear microscopy positivity (4/9, 44.4%) with fewer graded as scanty or 1+ (1/4, 25.0%); a higher rate of Xpert Ultra positivity (9/9, 100%) with fewer graded semi-quantitatively as “trace” or “very low” (3/9, 33.3%); and shorter MGIT culture time-to-positivity (12 days, IQR 8–18). However, there were few symptomatic TB cases and thus differences compared to asymptomatic TB were not formally tested.

Seven asymptomatic TB cases (7/42, 16.7%) were diagnosed by Xpert Ultra alone (MGIT culture negative), only two of whom had a previous TB episode within the prior three years (1.4 and 1.8 years). There were also three asymptomatic HHC with negative MGIT culture and trace positive Xpert Ultra results (not included in the MRS); 2/3 (33.3%) of whom had a known prior TB episode within the preceding three years.

Fifteen of the 18 (83.3%) asymptomatic TB cases without CXR abnormality suggestive of TB were sputum MGIT culture positive, with longer median MGIT culture time-to-positivity of 16 days (IQR 10–19), very low Xpert Ultra semi-quantitative *Mtb* load (5/15 negative, 6/15 “trace” or “very low”, and 2/15 “low”, 2/15 “medium” or “high”), and 11/15 (73.3%) were smear negative. The three MGIT culture negative asymptomatic CXR negative participants all had “very low” semi-quantitative Xpert Ultra results, and negative sputum smears; Two of the three had prior TB episodes, both more than three years prior.

### Incremental yield of screening and number needed to test to diagnose one case of TB

Universal sputum microbiological testing detected 18 asymptomatic TB cases (18/51; sensitivity 35.3%, 95%CI 23.6–49.0%) that would have been missed by symptom- and CXR-triggered investigation for TB (**Figure 3**), increasing yield by 54.5% (18/33; 95%CI 38.0–70.2%).

**Figure 3.**
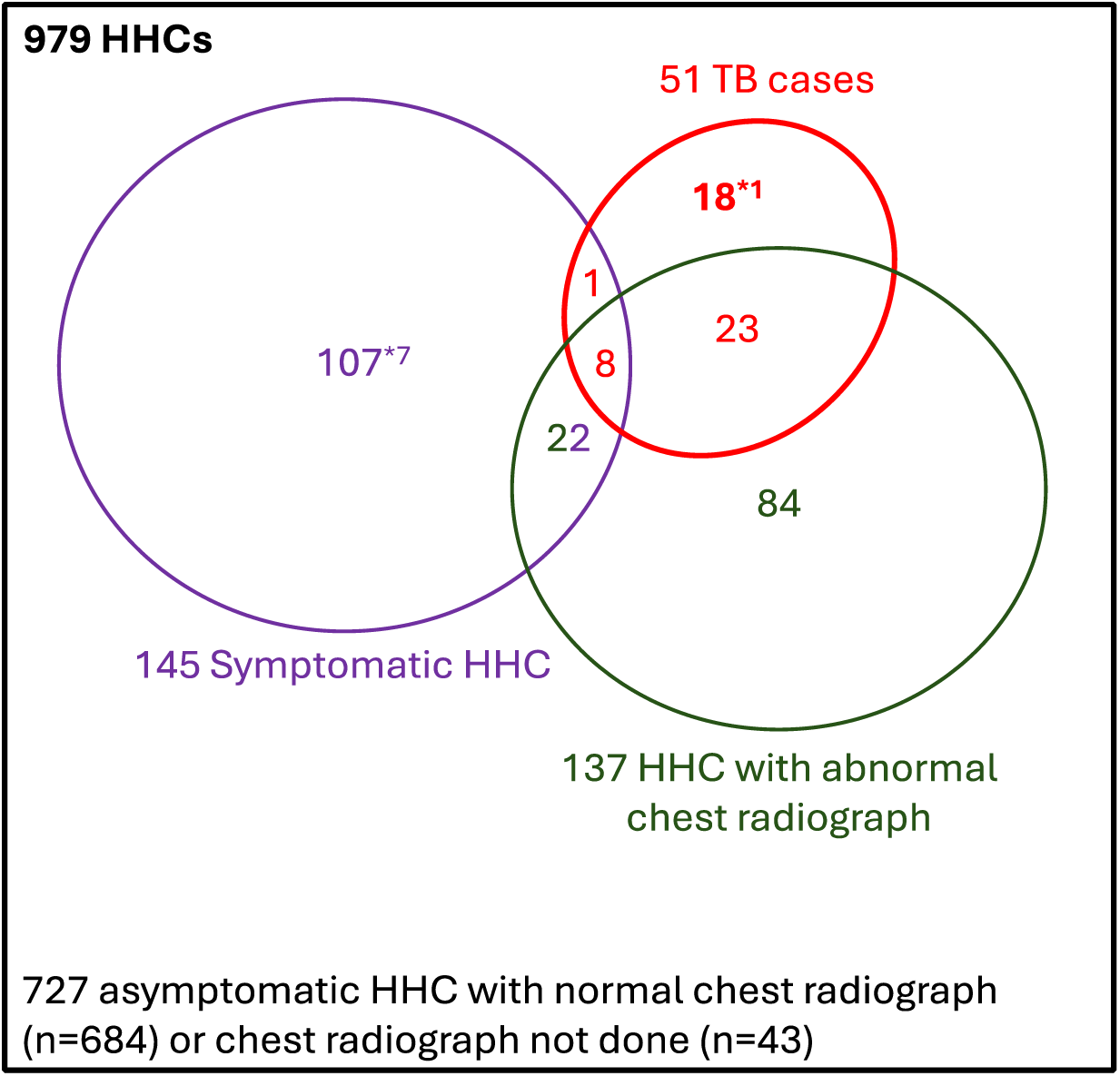
Venn diagram demonstrating yield of symptom screening and chest radiography (CXR) for detection of pulmonary tuberculosis (TB) among household contacts (HHC). *There were 7 symptomatic HHC without TB and 1 asymptomatic HHC with TB who did not have CXR.

Universal testing of HHC with both sputum Xpert Ultra and MGIT culture (MRS) required 19.2 (95%CI 14.7–25.1) confirmatory tests to diagnose one case of TB. We explored the incremental yield (sensitivity) and number of confirmatory tests needed to diagnose one TB case for different screening strategies (**Table 2**). A symptom screening approach with sensitivity 17.6% (95%CI 9.6– 30.3%) would have missed 42/51 TB cases (82.4%, 95%CI 69.7–90.4%). CXR screening with sensitivity 62.0% (95%CI 48.2–74.1) would have missed 19/50 (38.0%, 95%CI 25.9–51.8%) of all TB cases, but reduced the number of confirmatory tests needed to diagnose one TB case to 7.7 (5.6–10.6) with, or 4.4 (3.3–6.1) without, parallel symptom screening, respectively. The addition of symptom screening in parallel to CXR screening marginally increased sensitivity to 64.0% (95%CI 50.1–75.9%). However, among asymptomatic HHC, CXR screening with sensitivity 56.1% (95%CI 41.0–70.1%) would have missed 18/41 (43.9%, 95%CI 29.9–59.0%) of asymptomatic TB cases.

**Table 2.** Comparison of yield and number needed to test of different TB screening approaches for detection of TB cases among household contacts of index TB cases.

| Initial screening approach | Number of participants included in analysis | Number of positive screening tests (%) | Number of TB cases diagnosed (sensitivity, 95% CI)<sup>a</sup> | NNT with confirmatory tests to diagnose 1 case (95% CI) |
| --- | --- | --- | --- | --- |
| No screening; MRS for all<sup>a</sup> | 979 | 979 (100%) | 51/51<br>(100%, 93.0–100) | 19.2 (14.7–25.1) |
| Symptom screening<sup>b</sup> | 979 | 145 (14.8%) | 9/51<br>(17.6%, 9.6–30.3) | 16.1 (8.8–30.3) |
| CXR screening<sup>c</sup> | 929<sup>e</sup> | 137 (14.7%) | 31/50<br>(62.0%, 48.2–74.1) | 4.4 (3.3–6.1) |
| Parallel symptom and CXR screening<sup>d</sup> | 929<sup>e</sup> | 245 (26.4%) | 32/50<br>(64.0%, 50.1–75.9) | 7.7 (5.6–10.6) |
| CXR screening among asymptomatic participants | 791<sup>f</sup> | 107 (13.5%) | 23/41<br>(56.1%, 41.0–70.1) | 4.7 (3.3–6.8) |

<sup>a</sup> Microbiological reference standard (MRS) is positive sputum culture or Xpert Ultra, excluding Trace positive results.
<sup>b</sup> Any symptom of any duration.
<sup>c</sup> Any chest radiography (CXR) abnormality.
<sup>d</sup> Any symptom of any duration or any CXR abnormality.
<sup>e</sup> 50 participants without CXR, including 1 asymptomatic TB case, were excluded.
<sup>f</sup> 43 asymptomatic participants without CXR, including 1 asymptomatic TB case, were excluded.
NNT, number needed to test.

### C-reactive protein has low accuracy for TB screening among exposed household contacts

CRP was measured in a subset of 25 asymptomatic and 7 symptomatic pulmonary TB cases and 126 healthy HHC from this cohort; and 607 symptomatic adults presenting to clinic with TB symptoms (199 symptomatic pulmonary TB cases and 408 symptomatic controls without TB). CRP concentrations appeared higher among symptomatic TB cases (median 17.8 mg/dL, IQR 8.1−29.4) presenting for healthcare than in symptomatic TB cases detected among HHC (median 4.3 mg/dL, IQR 1.7−11.0; **Figure 4**).

CRP was able to differentiate symptomatic clinic attendees with and without symptomatic TB (AUC 0.84, 95%CI 0.81–0.88), and HHC with and without symptomatic TB (AUC 0.74, 95%CI 0.52−0.96), but was not able to discriminate between HHC with asymptomatic TB disease from those without TB (AUC 0.60, 95%CI 0.47–0.73). Sensitivity of CRP for diagnosing asymptomatic TB among HHC was 48.0% (95%CI 30.0–66.5%) when specificity was set at 70% per WHO Target Product Profile minimum screening test criteria^13^. Conversely, with sensitivity set at 90%, specificity was 19.0% (95%CI 13.1–26.8%).

**Figure 4.**
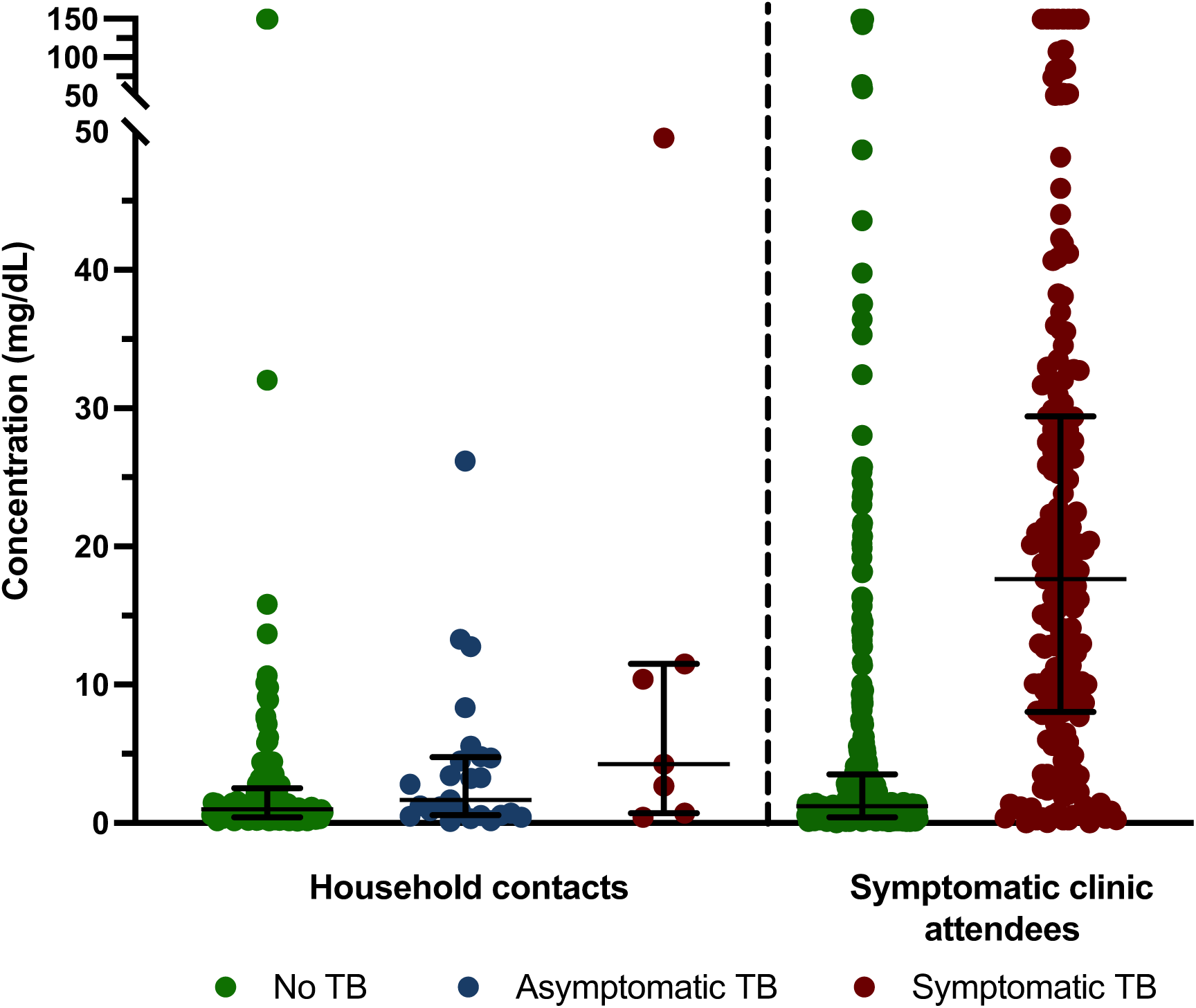
C-reactive protein (CRP) is unable to differentiate asymptomatic tuberculosis (TB) cases from asymptomatic household contacts (HHC) without TB. CRP concentration measured by multiplex bead array in a subset of 25 asymptomatic and 7 symptomatic pulmonary TB cases, and 126 healthy HHC, from this cohort, and 607 symptomatic adults presenting to clinic with TB symptoms (199 symptomatic pulmonary TB and 408 without TB). CRP concentrations were censored at 150 mg/dL. Each dot represents one participant. Boxes depict the IQR, the midline represents the median (shown), and the whiskers indicate the IQR ± (1.5 × IQR).

## DISCUSSION

We aimed to understand the burden and clinical characteristics of asymptomatic TB among TB-exposed HHC in three communities in South Africa, and to measure the yield of universal symptom, CXR, and sputum screening. We found 5.2% TB prevalence among HHC, which is more than 6-fold higher than the community rate reported in the South African national TB prevalence survey.^2^ We have shown that reliance on symptom screening to trigger diagnostic TB investigation would have missed 82.4% of microbiologically-confirmed, asymptomatic TB cases in this cohort, in keeping with similar studies conducted in the community.^14^ The yield of asymptomatic TB cases (n=42; 82.4%) detected by active screening of all exposed HHC, regardless of presence of symptoms, was 4.7-fold that of symptomatic TB cases (n=9; 17.6%). This ratio is consistent with modelling estimates of household *Mtb* exposure and infection occurring within the proceeding 6–12 months.^15^

The yield of CXR screening for all TB, alone or in combination with symptom screening, was low — 62.0% and 64.0%, respectively. The observed sensitivity of CXR screening for all TB among HHC was considerably lower than the 94.7% expected by WHO, but is consistent with the pooled 62.4% sensitivity derived from review of community-based screening studies (**Figure 1**). It is also notable that 43.9% of asymptomatic TB cases with normal CXR would have been missed by traditional screening methods. Stuck and colleagues modelled the prevalence of asymptomatic pulmonary TB in adults in community settings, and estimated even lower CXR sensitivity of 25% among individuals with no cough.^16^

The universal TB sputum screening methodology employed in this study detected 54.5% more TB cases than would a parallel symptom- and CXR-triggered approach, such as that adopted by the South African national TB prevalence survey.^2^ Extrapolating our findings among HHC in three TB-endemic communities to the South Africa-wide population survey, which observed a microbiologically-confirmed pulmonary TB prevalence of 852 cases (95%CI 679–1026) per 100 000 population, the true prevalence of microbiologically-confirmed asymptomatic TB cases, including CXR-negative cases, might be as high as 1,331 (95%CI 1061–1603) per 100 000 population. If these findings from South Africa are replicated in other high TB burden countries in which national prevalence surveys omitted universal sputum microbiological testing of symptom- and chest radiograph-negative persons, it is likely that the global burden of asymptomatic TB has been significantly under-estimated. The screening approach also has implications for TB control. The TREATS and SCALE community-based, cluster-randomised studies^17,18^ used symptom and CXR screening, but failed to show impact on TB incidence.^19^ Conversely, the ACT3 Study in Vietnam performed universal TB sputum Xpert testing and demonstrated a reduction in incidence.^19,20^ Incomplete treatment of asymptomatic TB cases in the absence of universal sputum testing might explain this difference in outcome.

Our finding that symptom- and CXR-screening tools, developed for triage of symptomatic TB patients seeking care, perform poorly as screening tests for asymptomatic TB in the community, is mirrored by our findings for CRP, which has recently been recommended by the WHO as a TB screening tool for people living with HIV.^11^ Recent studies have demonstrated promising performance of CRP for TB triage among symptomatic adults and individuals attending outpatient clinics.^21,22^ However, in our exploratory analysis, serum CRP levels were lower among TB cases among HHC in the community compared to TB cases presenting to clinics, in keeping with the study by Kendall and colleagues.^23^ Our findings also mirror those of Ruperez et al;^24^ CRP did not significantly differentiate asymptomatic TB from asymptomatic HHC without TB, suggesting that CRP would not be appropriate as a screening test in a community setting.

Asymptomatic TB cases also had predominantly paucibacillary sputum, with low Xpert Ultra semi-quantitative and sputum smear grade, and low CRP levels. Collectively, these data support the hypothesis that asymptomatic TB in the community, among persons not presenting for healthcare, is associated with less severe disease and less systemic inflammation, than symptomatic TB diagnosed among clinic attendees. This finding reinforces the principle that a triage test for symptomatic clinic attendees cannot be adopted uncritically as a community screening test among asymptomatic individuals. It will be essential that performance of new screening tests for asymptomatic TB in the community is not benchmarked against performance of those same tests for triage and diagnosis of symptomatic TB. Further, given the importance of mass community screening to detect asymptomatic TB, it has been proposed that tests with sensitivity lower than proposed in the WHO Target Product Profile^13^ might be acceptable.^25^

It is acknowledged that mass community CXR screening in endemic settings may detect a large number of undiagnosed TB cases if deployed at scale^26^, despite suboptimal sensitivity. However, control of the epidemic may require that a larger proportion of asymptomatic community TB cases are detected and treated, in which case, additional diagnostic tools would be needed to find the approximately 40% of TB cases missed by CXR screening of asymptomatic individuals. In the absence of a more sensitive screening tool for asymptomatic TB, our study suggests that universal sputum testing with either a molecular test or culture is appropriate for selected high-risk groups, such as HHC, with high diagnostic yield. In community prevalence surveys, the yield of prevalent TB is low (<0.1–1.2%), with asymptomatic TB estimated at around 50% of cases.^3^ In an individual participant data meta-analysis, Stuck et al. (2024)^16^ reported a lower proportion of asymptomatic pulmonary TB in adults in community settings of 28% after adjustment. Our study indicates that an additional 36% of asymptomatic TB cases could be accrued through universal sputum testing, but this approach might not be affordable in many high TB burden countries.

A particular strength of this study is the systematic approach to universal symptom, CXR, and sputum screening. There were also several limitations. Only spontaneously expectorated sputum samples were collected; it is possible that sputum induction may have yielded higher rates of asymptomatic TB. We detected eight sputum MGIT culture negative, Xpert Ultra positive TB cases, five of whom had prior TB. However, only two of the prior episodes were within the preceding three years, which makes false positive Xpert results less likely. It is also possible that the CXR reading methodology may have missed pathology. CXR were read once by an investigator at each site for any abnormality and, while it is possible that additional readers might increase sensitivity, it is more likely that requiring concordance of two or more readers would improve specificity at the expense of sensitivity.^27^ Specialised radiological training and expertise might further increase accuracy, but may not be readily available in TB-endemic settings. CXR reading with computer-assisted detection (CAD) software meets or exceeds the minimum TPP for a TB triage test for symptomatic adults with presumptive TB,^13,22,28^ and has been recommended by the WHO as an alternative to human readers. However, in the South African TB prevalence survey CAD accuracy was significantly lower in asymptomatic compared to symptomatic individuals.^29^ In future work, we plan to compare diagnostic accuracy of CXR with multiple investigator readers to CAD for diagnosis of asymptomatic TB. CRP levels were measured by multiplex bead array, rather than a validated, high-sensitivity assay platform. However, although assay sensitivity might differ, it appears that relative to symptomatic TB in symptomatic clinic attendees, CRP levels were lower in HHC with asymptomatic or symptomatic TB. Finally, TB symptoms were self-reported and subjective. Chronic or mild symptoms that did not interfere with daily function, such as a smoking-related cough, may have been perceived as normal. Perhaps more important than whether symptoms were truly present or absent, unlike a clinic triage scenario, all HHC who participated in this study were not actively seeking healthcare.

Our study supports the idea that current community screening approaches detect only the “tip of the iceberg”^30^, since symptom and CXR screening missed approximately 40% of asymptomatic TB in this household contact-tracing study. If these findings in South Africa are replicated in other high burden countries, in which national TB prevalence surveys have omitted definitive diagnostic testing of asymptomatic, CXR-negative individuals, it is possible that the global burden of asymptomatic TB has been significantly underestimated. It also appears that biomarkers developed and tested for triage of symptomatic TB, such as CRP, would perform sub-optimally as community screening tests for asymptomatic TB. These findings suggest that, pending discovery and validation of new non-sputum TB biomarkers for asymptomatic TB, accurate community-based TB screening requires universal sputum microbiological testing.

## AUTHORS’ CONTRIBUTIONS

TJS, TRS, and MH conceived the idea, raised funds, and/or provided the resources. GW, TJS, TRS, and MH wrote the study protocol and provided study oversight. MT, TM, STM, and FN and were responsible for all site-level activities, including recruitment, clinical management, and data collection. KH provided operational support, and project management. NNC performed Luminex bead arrays. HM, AK, FM, RP, KH, KS, and YFvdH cleaned and verified the underlying data. SCM, HM, and DoA analysed the data. SCM, HM, and MH interpreted the results and wrote the first draft. All authors had full access to the data, confirm the integrity of the data and its presentation, agree with its interpretation as discussed in the manuscript, and reviewed, revised, and approved the manuscript before submission. The corresponding author had final responsibility for the decision to submit for publication.

## DATA AVAILABILITY STATEMENT

The data that support the findings of this study are available on request from the corresponding author. The data are not publicly available due to international data privacy regulations and ethical restrictions.

