## Supplement for "Screening for Asymptomatic Tuberculosis among Adults with Household Exposure to a Patient with Pulmonary Tuberculosis"

### RePORT South Africa Study Team

| Name | Affiliation |
|---|---|
| Pattamukkil Abraham | Perinatal HIV Research Unit, University of Witwatersrand |
| Denis Awany | South African Tuberculosis Vaccine Initiative, University of Cape Town |
| Cynthia Baard | University of Cape Town Lung Institute |
| Zainab Baig | Africa Health Research Institute |
| John Belisle | Colorado State University |
| Rebecca Berhanu | Vanderbilt Tuberculosis Center, Vanderbilt University |
| Nicole Bilek | South African Tuberculosis Vaccine Initiative, University of Cape Town |
| Gerard Cangelosi | University of Washington |
| ACE Carstens | Department of Biomedical Sciences, Stellenbosch University |
| Kevyna Chetty | Africa Health Research Institute |
| Yolundi Cloete | South African Tuberculosis Vaccine Initiative, University of Cape Town |
| Rodney Dawson | University of Cape Town Lung Institute |
| Marwou de Kock | South African Tuberculosis Vaccine Initiative, University of Cape Town |
| Keertan Dheda | University of Cape Town Lung Institute |
| Gareta Dickman | Africa Health Research Institute |
| Charity Dire | Perinatal HIV Research Unit, University of Witwatersrand |
| Karen Dobos | Colorado State University |
| Stephany Norah Duda | Vanderbilt Tuberculosis Center, Vanderbilt University |
| Mzwandile Erasmus | South African Tuberculosis Vaccine Initiative, University of Cape Town |
| Aliasgar Esmail | Division of Pulmonology, Department of Medicine, Groote Schuur Hospital and University of Cape Town Lung Institute |
| Marina Cruvinel Figueiredo | Vanderbilt Tuberculosis Center, Vanderbilt University |
| Marika Flinn | Department of Biomedical Sciences, Stellenbosch University |
| Bernard Fourie | University of Pretoria |
| Travis Harris | Vanderbilt Tuberculosis Center, Vanderbilt University |
| Andriette Hiemstra | Department of Biomedical Sciences, Stellenbosch University |
| Shameem Jaumdally | University of Cape Town Lung Institute |
| Ryan Johnson | University of Cape Town |
| Farina Karim | Africa Health Research Institute |
| Masooda Kaskar | South African Tuberculosis Vaccine Initiative, University of Cape Town |
| Nobulumko Khomba | South African Tuberculosis Vaccine Initiative, University of Cape Town |
| Thandeka Khoza | Africa Health Research Institute |
| Léanie Kleynhans | Department of Biomedical Sciences, Stellenbosch University; Mater Research Institute – The University of Queensland |
| Tahira Kootbodien | University of Cape Town |
| Andrea Kotze | University of Cape Town Lung Institute |
| Belinda A Kriel | Department of Biomedical Sciences, Stellenbosch University |
| Ané Kruger | Department of Biomedical Sciences, Stellenbosch University |
| Alasdair Leslie | Africa Health Research Institute |
| Lorraine Lichakane | Perinatal HIV Research Unit, University of Witwatersrand |
| Ilze Louw | Department of Biomedical Sciences, Stellenbosch University |
| Angelique Luabeya | South African Tuberculosis Vaccine Initiative, University of Cape Town |
| Simbarashe Mabwe | South African Tuberculosis Vaccine Initiative, University of Cape Town |
| Candice MacDonald | Department of Biomedical Sciences, Stellenbosch University |
| Lindiwe Madziwa | Africa Health Research Institute |
| Lebohang Makhethe | South African Tuberculosis Vaccine Initiative, University of Cape Town |
| Sandisiwe Mangali | South African Tuberculosis Vaccine Initiative, University of Cape Town |
| Neil Martinson | Perinatal HIV Research Unit, University of Witwatersrand |
| Linda Mbuthini | University of Cape Town |
| Carolina Mehaffy | Colorado State University |
| Thabang Moloja | Perinatal HIV Research Unit, University of Witwatersrand |
| Angelique Mouton | South African Tuberculosis Vaccine Initiative, University of Cape Town |
| Mbusiseni Ngema | Perinatal HIV Research Unit, University of Witwatersrand |

| | |
|---|---|
| Hlengiwe Nkambule | South African Tuberculosis Vaccine Initiative, University of Cape Town |
| Onke Nombida | South African Tuberculosis Vaccine Initiative, University of Cape Town |
| Sarah Nyangu | South African Tuberculosis Vaccine Initiative, University of Cape Town |
| Fajwa Opperman | South African Tuberculosis Vaccine Initiative, University of Cape Town |
| Gregory Ording-Jespersen | Africa Health Research Institute |
| Kennedy Otwombe | Perinatal HIV Research Unit, University of Witwatersrand |
| Tahlia Perumal | University of Cape Town Lung Institute |
| Tracy Richardson | Department of Biomedical Sciences, Stellenbosch University |
| Carmen Segelaar | South African Tuberculosis Vaccine Initiative, University of Cape Town |
| Jane Shaw | Department of Biomedical Sciences, Stellenbosch University |
| Kimberly Shelton | Colorado State University |
| Justin Shenje | South African Tuberculosis Vaccine Initiative, University of Cape Town |
| Theresa Smit | Africa Health Research Institute |
| Bronwyn Smith | Department of Biomedical Sciences, Stellenbosch University |
| Candice Snyders | Department of Biomedical Sciences, Stellenbosch University |
| Marcia Steyn | South African Tuberculosis Vaccine Initiative, University of Cape Town |
| Sara Suliman | University of California San Francisco |
| Floris Swanepoel | Perinatal HIV Research Unit, University of Witwatersrand |
| Lorraine Thobakgale | Perinatal HIV Research Unit, University of Witwatersrand |
| Susanne Tonsing | Department of Biomedical Sciences, Stellenbosch University |
| Nicolette Tredoux | South African Tuberculosis Vaccine Initiative, University of Cape Town |
| Megan Turner | Vanderbilt Tuberculosis Center, Vanderbilt University |
| Petrus Tyambetyu | South African Tuberculosis Vaccine Initiative, University of Cape Town |
| Habibullah Valley | South African Tuberculosis Vaccine Initiative, University of Cape Town |
| Lyle van de Berg | Perinatal HIV Research Unit, University of Witwatersrand |
| Gian van der Spuy | Department of Biomedical Sciences, Stellenbosch University |
| Ilana R van Rensburg | Department of Biomedical Sciences, Stellenbosch University |
| Johanna E van Rooyen | South African Tuberculosis Vaccine Initiative, University of Cape Town |
| Hilary Vansell Riley | Vanderbilt Tuberculosis Center, Vanderbilt University |
| Ashley Veldsman | South African Tuberculosis Vaccine Initiative, University of Cape Town |
| Lindsay Wilson | University of Cape Town |
| Rachel C Wood | University of Washington |
| Lesley Workman | University of Cape Town |
| Heather Zar | Department of Paediatrics and Child Health, University of Cape Town |

### RePORT South Africa Study Design

The Regional Prospective Observational Research in Tuberculosis (RePORT) South Africa network recruits into two contemporaneous cohorts at seven research sites in South Africa. Cohort A recruits symptomatic clinic attendees with suspected TB to evaluate triage tests for symptomatic TB disease (passive case-finding); Cohort B recruits household or other close contacts of TB patients in the community to evaluate screening tests for both symptomatic and asymptomatic TB disease (active case-finding).

**Cohort A:** Adults (≥18 years) presenting to healthcare clinics with presumptive pulmonary TB were enrolled at five sites in South Africa (Ravensmead, Klipfontein, and Khayelitsha, Western Cape Province; Durban, KwaZulu-Natal Province; and Pretoria, Gauteng Province). Participants reported at least one symptom consistent with pulmonary TB, including cough, fever, weight loss, fatigue, night sweats, pleuritic chest pain, or haemoptysis, for any duration. Participants were excluded if they were unlikely to attend follow-up visits, had active psychiatric conditions or alcohol/drug dependence interfering with study adherence, or had received anti-TB treatment for more than 7 days within 30 days prior to screening.

Spontaneously expectorated sputum collection for Xpert Ultra (Cepheid, CA, USA) and liquid culture (Mycobacteria Growth Indicator Tube [MGIT], BACTEC, Beckton Dickinson, NJ, USA) was performed at baseline for all participants. The microbiological reference standard (MRS) for microbiologically-confirmed pulmonary TB disease was at least one sputum specimen positive by MGIT culture or Xpert Ultra (excluding trace positive results). Sputum induction was not performed. Participants who were unproductive of sputum, or with Xpert Ultra trace positive result only, were considered sputum-negative in the primary analysis. Participants who, after investigation for TB at baseline, were not confirmed by positive sputum MGIT culture or Xpert Ultra, were defined as controls for the purpose of diagnostic analyses. The definition of prevalent TB was restricted to cases diagnosed on sputum samples collected at baseline. All participants diagnosed with TB disease were referred for treatment.

### Studies Included in Systematic Review, by Year of Publication
